# Exploratory Biomarkers for Acute Rejection in Vascularized Composite Allotransplantation

**DOI:** 10.1101/2025.07.22.25331528

**Authors:** Dominika Pullmann, William J Rifkin, Haruyuki Hirayama, Bruce E Gelb, Ata S Moshiri, Massimo Mangiola, Eduardo D Rodriguez, Catherine P Lu, Piul S Rabbani

## Abstract

Vascularized composite allotransplantation (VCA) involves immunologically heterogeneous tissues and is associated with high rates of acute rejection (AR), particularly due to the immunogenicity of the skin. This study assessed biomarkers to monitor AR in a recipient of full-face and bilateral hand transplantation. Over 4.6 years postoperatively, we analyzed serial blood and tissue samples, including donor-derived cell-free DNA (dd-cfDNA) in plasma, recipient DNA in allograft biopsies using short tandem repeat (STR) analysis, lymphocyte subsets via flow cytometry, and level of angiotensin II type 1 receptor antibodies (AT1R-Abs). dd-cfDNA and STR showed trends toward elevated recipient signal during acute rejection, though differences were not statistically significant. CD8+ T-cell percentages increased before AR onset, suggesting potential as a prognostic biomarker. AT1R-Ab levels did not differ significantly between AR and non-rejection episodes, possibly due to prophylactic immune cell depletion. While dd-cfDNA and STR levels correlate with rejection episodes and reflect key cellular events within graft tissue, CD8+ T-cell count remains the most robust prognostic biomarker for predicting the onset of cytotoxicity in this patient. These findings highlight the importance of further longitudinal, multi-patient studies to validate emerging biomarkers and improve rejection monitoring strategies in VCA.

## Introduction

Vascularized Composite Allotransplantation (VCA) involves concurrent transplantation of heterogenous organs –skin, muscle, bone, nerves and associated vasculature. Among these, skin is the most immunogenic component contributing to VCA’s rejection rate, which is 7-fold higher than that of solid organ transplants (SOT).^1^ Despite immunosuppressive regimens, up to 85% of VCA recipients experience acute rejection (AR), potentially progressing to chronic rejection (CR) and graft failure.^2^ Banff criteria-based histopathological evaluation of VCA skin, adapted from SOT, is the gold standard for assessing rejection, but is limited by sampling variability, interpretive subjectivity, local inflammation, and cosmetic injury.^3^ Non-invasive imaging, like MRI and ultrasonography, shows promise for detecting rejection, but lacks resolution for subtle changes and early vasculopathy.^4^ Evaluating alternative biomarkers for timely and sensitive VCA AR monitoring is crucial for clinical management.

Unlike SOT, VCA’s constituent organs have varying immunogenic profiles, complicating rejection detection. Rejection can occur without visible signs in deeper tissues, leading to insidious graft deterioration. VCA lacks validated biomarkers that correlate with rejection severity or predict AR episodes before clinical manifestation. Some VCA studies report skin redness and edema as early signs, with CD8^+^-cytotoxicity contributing to keratinocyte exocytosis; however, the relative contribution of donor vs recipient T-cells is unclear.^1^ In contrast, Short Tandem Repeats (STR) profiling, used in solid/hematologic transplants, analyzes the ratio of recipient-to-donor DNA using their respective unique single nucleotide polymorphisms (SNP) in the donor graft tissue.^5,6^ In turn, the ratio of donor-derived-cell-free DNA (dd-cfDNA) to total cfDNA in recipient blood can reveal graft cellular injury/death.

SOT studies show detectable elevation in plasma dd-cfDNA earlier than confirmatory/clinical AR diagnoses.^7–9^ Both SOT and VCA research have identified non-HLA-associated serum antibodies such as anti-angiotensin-II-type-1-receptor antibodies (AT1R-Abs) as promising rejection biomarkers.^10–12^ Identifying comprehensive biomarkers that capture the status of the entire VCA is critical and warrants further investigation and innovation. Here, we investigated the association between these markers and AR in a VCA recipient.

## Material and Methods

### Patient

We analyzed data from a male patient in his 20’s, full face and bilateral hand allograft recipient, approved by the IRB at NYU. Along with clinical appearance and symptoms, dermatopathological evaluation of H&E skin biopsies using Banff grading for immune cell infiltration and epidermal-dermal-junction changes^13^ defined rejection episodes. The patient received intravenous steroid therapy, plasmapheresis with exchange, and belatacept infusions for rejection episodes (Fig. 1B).

**Figure 1.**
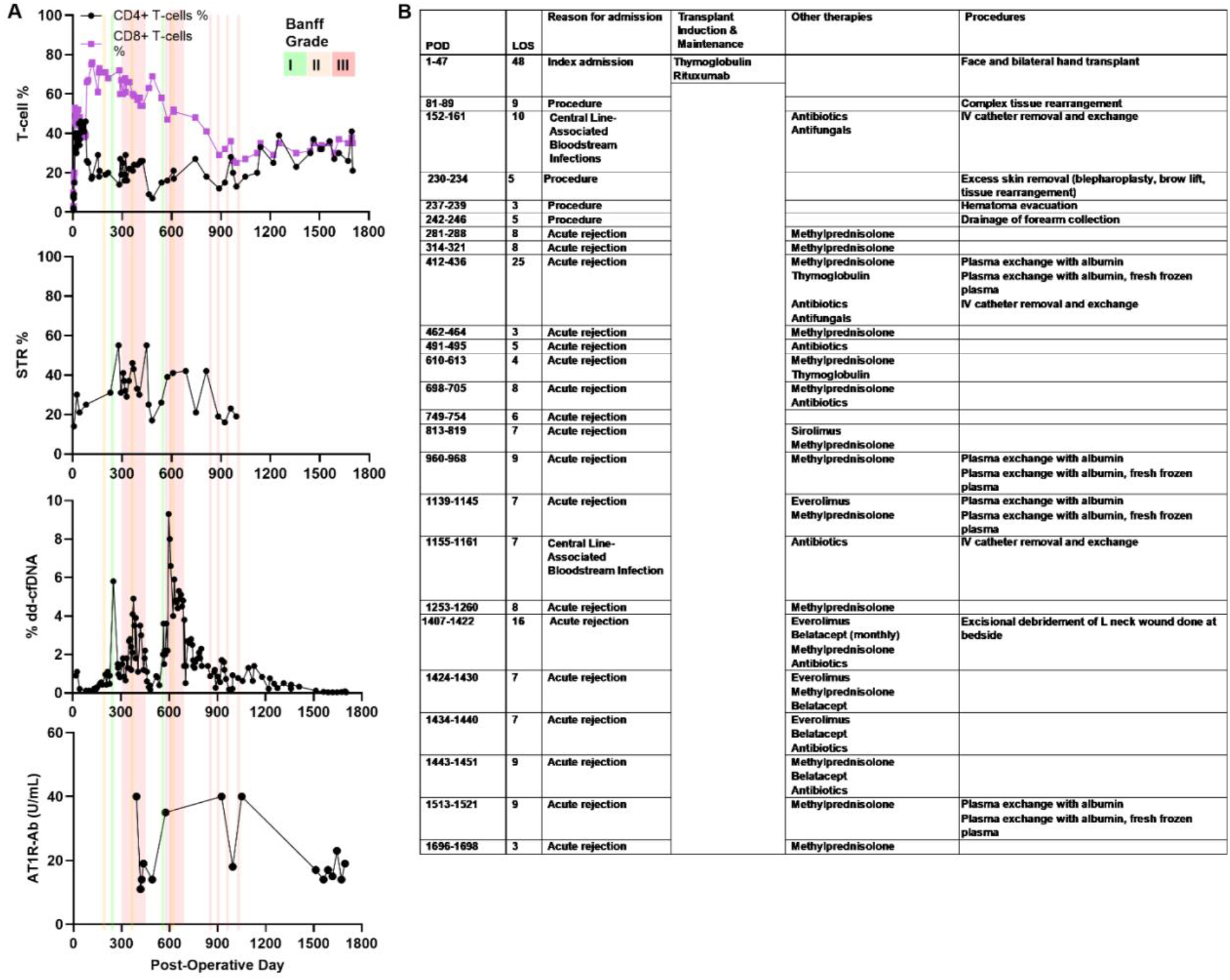
Post-operative rejection monitoring and management. **A.** Rejection monitoring markers and Banff grades. The lack of Banff data after POD 900 reflects the shift from a biopsy-based diagnosis to a holistic clinical assessment of AR. **B**. Summary of treatments during inpatient admissions for revision procedures or AR.

### Blood and Allograft Sampling

We evaluated samples collected over 4.6 years, up to post-operative day (POD) 1700 (Fig. 1A). We analyzed 83 blood samples for lymphocyte subsets by flow cytometry, 30 allograft skin biopsies for STR, 121 plasma samples for dd-cfDNA (Allosure, CareDx, CA), and 16 serum samples for AT1R-Abs detection (UCLA Immunogenetics Laboratory, CA) using ELISA (EIA-AT1RX, ThermoFisher, MA).

### Statistical Analysis

We considered samples collected before or after inpatient hospitalization for rejection as non-rejection (NR) data points. We used GraphPad Prism 9.02.0 for statistical analysis, p<0.05 being significant. We used the Shapiro-Wilk test for normality and a Wilcoxon nonparametric matched-pairs rank test to compare lymphocyte subsets, STR recipient DNA, dd-cfDNA, and AT1R-Ab during non-rejection and rejection episodes.

## Results

### Lymphocyte subsets

The median recipient CD3^+^CD4^+^-T-cells were 25% over the postoperative period (IQR 18-33%), 25% during non-rejection (IQR 18-35%, n=67), and 26% during rejection (IQR 17.25-29.75%, n=16; p=0.6741) (Fig. 2A). Median recipient CD3^+^CD8^+^-T-cells were 47% post-transplant (IQR 35-60%), 46% during non-rejection (IQR 32-58%, n=67), and 53% during rejection (IQR 36.5-62.5%, n=16; p=0.1509) (Fig. 2A).

**Figure 2.**
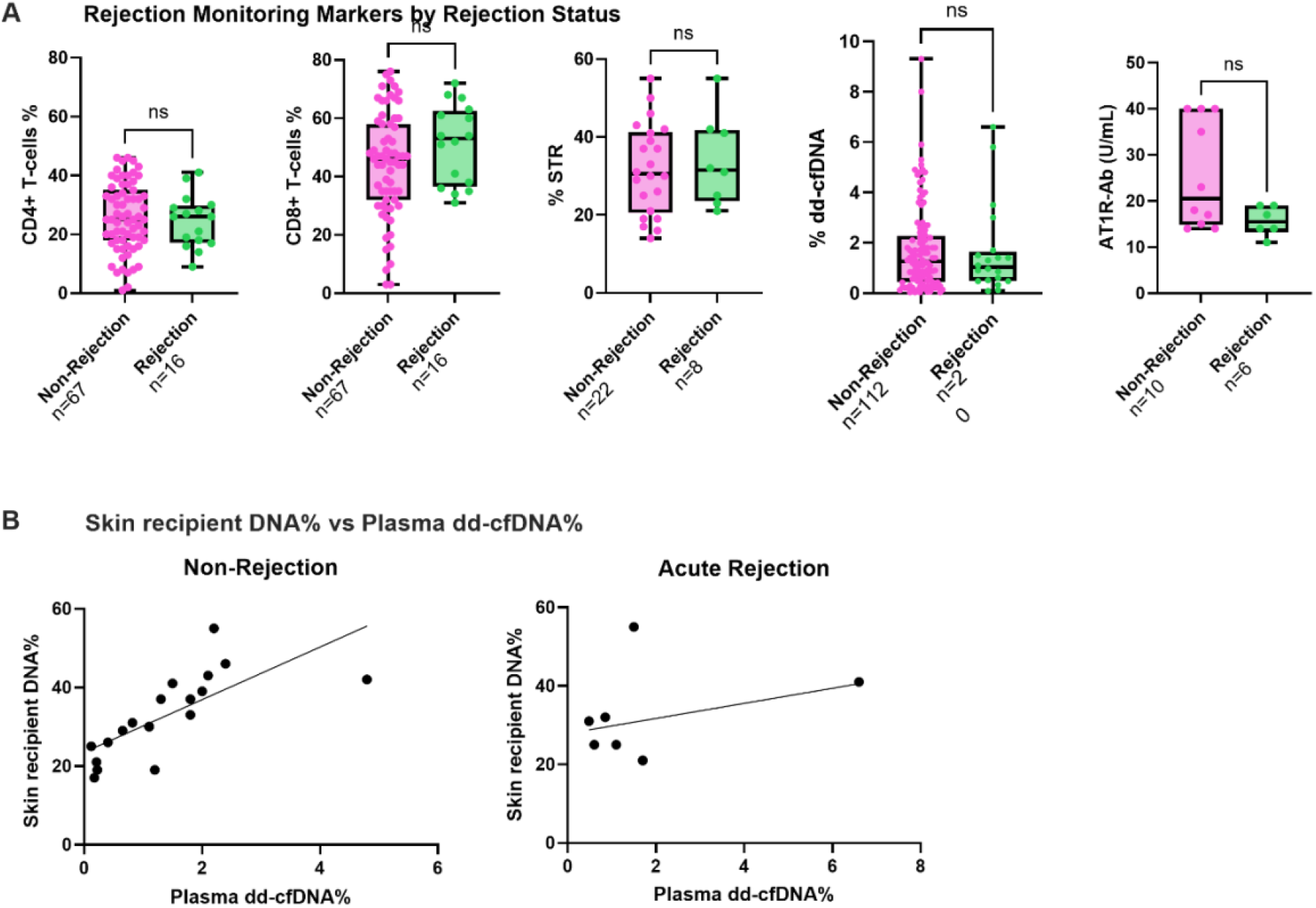
**A.** Rejection monitoring markers by rejection status. Box and whisker plots showing levels of five acute rejection markers: plasma percent dd-cfDNA (n=112 vs n=20), allograft skin % STR chimerism (n=22 vs n=8), CD4+ T-cell percentage (n=67 vs n=16), CD8+ T-cell percentage (n=67 vs n=16), and AT1R-Ab levels (n=10 vs n=6). Boxes show interquartile range; lines denote medians; whiskers indicate full range. **B**. Correlation between plasma and allograft skin dd-cfDNA% in paired samples. In NR samples (n=19), plasma dd-cfDNA and skin recipient DNA% showed a strong positive association (Spearman r=0.89, 95% CI: 0.73–0.96, p<0.0001). Simple linear regression yielded a significant model (y=6.69x+23.52) with R^2^=0.53 (p=0.0004), indicating a moderate linear relationship. In rejection samples (n=8), no significant association was observed between plasma and skin dd-cfDNA levels (Spearman r=-0.06, p=0.89). Linear regression analysis produced a non-significant model (y=1.92x+27.89) with R^2^=0.11 (p=0.43), suggesting poor predictive value of plasma dd-cfDNA for corresponding skin recipient DNA levels during AR.

Prior to POD900, subgroup analysis revealed similar recipient CD4^+^-T-cell percentages during non-rejection [25% (IQR 17-38%, n=51)] and AR [19% (IQR 16-26%, n=11; p=0.1237)], but a trend towards higher CD8^+^-T-cell percentages during AR [60% (IQR 52-67%, n=11) vs non-rejection 48% (IQR 44-61%, n=51; p=0.0587)]. CD8^+^-T-cells increased more than CD4^+^-T-cells relative to their respective baselines, 164 days before clinical rejection (76% on POD117 from 39% on POD77), suggesting predictive over diagnostic potential.

### Allograft skin STR

Overall median recipient DNA in allograft was 31% (IQR 22-41%, n=22), with no significant difference between non-rejection [30.5% (IQR 20.5-41.25%, n=8)] and AR [31.5% (IQR 23.5-41.75%; p=0.6872)] (Fig. 2A). STR analysis did not distinguish between non-rejection and rejection.

### Plasma dd-cfDNA

Although dd-cfDNA correlated with AR episodes (Fig. 1A), levels were similar between non-rejection [1.25% (IQR 0.45-2.275%, n=112)] and AR [1.035% (IQR 0.48-1.65%, n=20; p=0.6949)](Fig. 2A), with an overall median of 1.2% (IQR 0.48-2.2%, n=132). Despite this possible association, dd-cfDNA levels did not reliably distinguish between rejection and non-rejection.

### AT1R-Ab

Overall AT1R-Ab levels were 17.5% (IQR 14-32%), with no significant difference between non-rejection [20.5% (IQR 14.75-40%, n=10)] and AR [15.5% (IQR 13.25-19%, n=6; p=0.1206)] (Fig.2A); these results suggest AT1R-Ab levels do not distinguish between acute rejection and non-rejection.

## Discussion

This case study presents the longitudinal trends among exploratory surveillance biomarkers for rejection status in a VCA recipient such as T-lymphocyte subsets, allograft recipient DNA content, plasma dd-cfDNA levels, and AT1R-Abs. Our findings provide insight into the utility and limitations of these biomarkers in the context of VCA.

### Increased percentage of CD8^+^-T-cells precedes rejection

Consistent with literature, we found increased T-lymphocytes in rejection.^14^ Importantly, both CD4^+^ and CD8^+^-T-cell percentages nearly doubled by POD117, almost six months before the first AR episode (POD281), suggesting predictive potential. Subgroup analysis prior to POD900 suggested a trend toward higher CD8^+^-T-cell percentages during AR, consistent with their role in mediating cytotoxicity and graft injury.^14^ Further studies with a larger patient pool and frequent sampling may clarify the role of T-cell subsets in VCA rejection.

### Limited STR utility despite increased recipient DNA

STR analysis showed a non-statistically significant increase in recipient DNA within the allograft during AR, suggesting a potential, but still unclear, role for infiltrating recipient immune cells.^14^ However, clinical utility is limited by the need for biopsy causing graft injury and potential for sampling bias STR may not reflect the whole graft’s status.

### Detectable dd-cfDNA not discriminatory in VCA

Association between dd-cfDNA and AR was non-significant, but the trend aligns with its potential as a rejection marker, as in SOTs,^6–8,15^ but suggests lack of sensitivity/specificity in VCA. This could be due to complex tissue composition, processing of keratinocyte DNA by tissue macrophages, and highly immunogenic skin releasing dd-cfDNA under non-rejection stress (excess sun exposure, trauma, or infection).

### Limited utility of AT1R-Ab levels in a B-cell depleted patient

AT1R-Ab levels showed no significant differences between non-rejection and AR, contradicting their established role in rejection in SOTs.^10,11^ While preliminary, this result highlights limitations of AT1R-Ab as a biomarker, particularly when B/plasma cell is depleted as in our patient. This is among the first studies examining AT1R-Abs biomarker potential in VCA and warrants further investigation.

### Limitations and Future Directions

Our findings are limited by single-patient data and small sample size. The intermittent sampling schedule may not have captured AR-associated transient biomarker fluctuations. Successful immunosuppression resulted in NR sample-points superseding AR ones. Validating biomarkers need to address larger cohorts, frequent sampling and the evolving definition of VCA AR.

Despite limitations, these biomarkers offer potential for monitoring VCA graft health. Lymphocyte subsets, STR analysis, plasma dd-cfDNA and AT1R-Abs each provide unique insights into VCA status, though their individual diagnostic utility for AR remains unclear. Combining these biomarkers with advanced imaging or multi-omics approaches may improve diagnostic accuracy.

In summary, although we found no statistically significant differences in biomarker levels between non-rejection and AR, the observed trends warrant further research. Development and validation of biomarkers remains a critical priority for improving long-term outcomes for VCA recipients.

## Data Availability

All data produced in the present work are contained in the manuscript

## References

1. Leonard DA, Amin KR, Giele H, Fildes JE, Wong JKF. Skin Immunology and Rejection in VCA and Organ Transplantation. Curr Transplant Rep. 2020;7(4):251–259. doi:10.1007/s40472-020-00310-1

2. Petruzzo P, Lanzetta M, Dubernard JM, et al. The International Registry on Hand and Composite Tissue Transplantation. Transplantation. 2010;90(12):1590–1594. doi:10.1097/TP.0b013e3181ff1472

3. Krezdorn N, Lian CG, Wells M, et al. Chronic rejection of human face allografts. Am J Transplant Off J Am Soc Transplant Am Soc Transpl Surg. 2019;19(4):1168–1177. doi:10.1111/ajt.15143

4. Stead TS, Brydges HT, Laspro M, et al. Minimally and Non-invasive Approaches to Rejection Identification in Vascularized Composite Allotransplantation. Transplant Rev Orlando Fla. 2023;37(4):100790. doi:10.1016/j.trre.2023.100790

5. Cheng HY, Huang XT, Lin CF, et al. Predicting Outcomes of Rat Vascularized Composite Allotransplants through Quantitative Measurement of Chimerism with PCR-Amplified Short Tandem Repeat. J Immunol Res. 2020;2020:9243531. doi:10.1155/2020/9243531

6. Fernández-Galán E, Badenas C, Fondevila C, et al. Monitoring of Donor-Derived Cell-Free DNA by Short Tandem Repeats: Concentration of Total Cell-Free DNA and Fragment Size for Acute Rejection Risk Assessment in Liver Transplantation. Liver Transplant Off Publ Am Assoc Study Liver Dis Int Liver Transplant Soc. 2022;28(2):257–268. doi:10.1002/lt.26272

7. Stites E, Kumar D, Olaitan O, et al. High levels of dd-cfDNA identify patients with TCMR 1A and borderline allograft rejection at elevated risk of graft injury. Am J Transplant Off J Am Soc Transplant Am Soc Transpl Surg. 2020;20(9):2491–2498. doi:10.1111/ajt.15822

8. Huang E, Sethi S, Peng A, et al. Early clinical experience using donor-derived cell-free DNA to detect rejection in kidney transplant recipients. Am J Transplant Off J Am Soc Transplant Am Soc Transpl Surg. 2019;19(6):1663–1670. doi:10.1111/ajt.15289

9. Sindu D, Bay C, Grief K, Walia R, Tokman S. Clinical utility of plasma percent donor-derived cell-free DNA for lung allograft surveillance: A real-world single-center experience. JHLT Open. 2024;6:100141. doi:10.1016/j.jhlto.2024.100141

10. Dragun D, Müller DN, Bräsen JH, et al. Angiotensin II type 1-receptor activating antibodies in renal-allograft rejection. N Engl J Med. 2005;352(6):558–569. doi:10.1056/NEJMoa035717

11. Moreno JD, Verma AK, Kopecky BJ, et al. Angiotensin II Type 1 Receptor Antibody-mediated Rejection Following Orthotopic Heart Transplant: A Single-center Experience. Transplantation. 2022;106(2):373–380. doi:10.1097/TP.0000000000003712

12. Reinsmoen NL, Mirocha J, Ensor CR, et al. A 3-Center Study Reveals New Insights Into the Impact of Non-HLA Antibodies on Lung Transplantation Outcome. Transplantation. 2017;101(6):1215–1221. doi:10.1097/TP.0000000000001389

13. Cendales LC, Farris AB, Rosales I, et al. Banff 2022 Vascularized Composite Allotransplantation Meeting Report: Diagnostic criteria for vascular changes. Am J Transplant Off J Am Soc Transplant Am Soc Transpl Surg. 2024;24(5):716–723. doi:10.1016/j.ajt.2023.12.023

14. Frank R, Dean SA, Molina MR, Kamoun M, Lal P. Correlations of lymphocyte subset infiltrates with donor-specific antibodies and acute antibody-mediated rejection in endomyocardial biopsies. Cardiovasc Pathol Off J Soc Cardiovasc Pathol. 2015;24(3):168–172. doi:10.1016/j.carpath.2014.11.001

15. Teszak T, Barcziova T, Bödör C, et al. Donor-Derived Cell-Free DNA Versus Left Ventricular Longitudinal Strain and Strain-Derived Myocardial Work Indices for Identification of Heart Transplant Injury. Biomedicines. 2025;13(4):841. doi:10.3390/biomedicines13040841

